# Large Volume Micro-CT scanning of the dentoalveolar complex as a tool to evaluate dental health in an archaeological sample compared with traditional methods

**DOI:** 10.1101/2022.06.23.22276834

**Authors:** Angela Gurr, Denice Higgins, Maciej Henneberg, Jaliya Kumaratilake, Matthew Brook O’Donnell, Meghan McKinnon, Alan Henry Brook

## Abstract

**Background:** Archaeological investigations of human skeletal material require non-destructive techniques. Large Volume Micro-Computed Tomography (LV Micro-CT) scanning systems allow acquisition of data from complete skulls. This study aims to determine 1) whether LV Micro-CT scanning can as a single technique provide adequate data for the analysis of the dentoalveolar complex, 2) how its outputs of dental and alveolar bone analyses compare to those of macroscopic and radiographic methods, and 3) how it compares with Small Volume Micro-Computed Tomography (SV Micro-CT), for analysis of individual teeth?

**Material and Methods:** Five archaeological human skulls were investigated. These represented both sexes and a broad age range. Large Volume Micro-CT, Macroscopic and Radiographic methods and SV Micro-CT scanning were used. Statistical analysis of intra and inter-operator reproducibility using five observers was undertaken.

**Results:** Large Volume Micro-CT as a stand-alone technique provided results across the full range of dentoalveolar complex categories measured. By combining traditional techniques similar results were obtained. There were high levels of reproducibility for intra-operator scoring and good inter-operator agreement from 4 operators with 1 operator whose results were outliers.

**Discussion:** The LV Micro-CT was the *only* technique to singularly provide a full range of measurements. A combination of the other techniques covered a similar range of categories, but the use of multiple methods was more time consuming. For some specific measurements, SV Micro-CT provided more detailed information.

**Conclusion:** This first study confirms the value of LV Micro-CT scanning for the analysis of the dentoalveolar complex of archaeological samples. Findings demonstrate the advantage of LV Micro-CT, which provided a comprehensive range of data as a stand-alone technique rather than combing modalities to achieve the same result. The SV Micro-CT provided higher resolution analysis for loose individual teeth due to the small size of the specimens but could not study the alveolar bone.

## INTRODUCTION

Archaeological investigation of the human dentition and alveolar bone requires non-destructive analytical techniques. This applies to the rare skeletal sample of 19^th^ century settlers to South Australia, whose macro-skeletal findings have been published (1, 2). The Large Volume Micro-Computed Tomography (LV Micro-CT) scanning system is a promising non-destructive technological advance, which allows specimens that were previously too large to be micro-CT scanned, such as a human skull to be analysed at a high resolution.

Large Volume Micro-CT scanning has previously been used for clinical research (3-6), forensic investigations (7, 8), and some research of archaeological bone samples including the skeleton of King Richard III of England (9-13). However, none of these cases have focused on the dentoalveolar complex in-situ in the skull.

The currently available methods for the examination of the dentoalveolar complex in-situ in archaeological skull samples include macroscopic (visual) and radiographic techniques. Small Volume Micro-Computed Tomography (SV Micro-CT) scanning systems offers high-resolution data but can only accommodate small samples such as an individual tooth not in the jaw, and cannot provide joint data for teeth and the surrounding alveolar bone tissues. Therefore, to obtain the required information in relation to in-situ large specimens, such as the dentoalveolar complex, multiple techniques have to be employed, which can be time consuming and costly. Each of these methods provides different types of data. For example, an investigation of carious lesions on a tooth undertaken requires both macroscopic examination and dental radiographs (14).

This study aims to determine 1) if the LV Micro-CT scanning system as a single technique can provide adequate data for the analysis of the dental and alveolar bone tissue, 2) how its outputs compare to those of traditional macroscopic and radiographic methods, and 3) how LV Micro-CT compares to SV Micro-CT for the analysis of individual teeth?

## MATERIALS AND METHODS

### ARCHAEOLOGICAL SAMPLE

The skulls used in this investigation were from skeletal remains of individuals buried at the ‘free ground’ area of St Mary’s Anglican Church Cemetery, South Road, South Australia between 1847 and 1927 (1, 2). This area of the cemetery was unmarked and the 19^th^ century excavated individual skeletons cannot be identified. A site code and context number (e.g., St Mary’s Burial, number 73 = SMB 73) was assigned to each individual, as their burials were unmarked.

Well-preserved skulls of five individuals with dentitions in-situ were selected from both sexes and they represent a broad range of ages. The sex of the infant and subadults is undetermined due to the lack of morphological sexual differentiation in their skeletons. The selected individuals had the following tooth types in the in-situ dentoalveolar complexes available:

i. **SMB 73** - adult, male – age 35 - 45 years and had 19 permanent teeth – *Upper right*: central and lateral incisors (FDI notation system 11 & 12), canine (13) and the first molar (16). *Upper left*: central and lateral incisors (21 & 22), canine (23), second premolar (25) and the third molar (28). *Lower left*: central and lateral incisors (31, 32), canine (33), first and second premolar (34, 34). *Lower right*: central and lateral incisors (41, 42), canine (43), first and second premolars (44, 45).
ii. **SMB 66B** - adult, female - age 35 - 45 years and had 17 permanent teeth – *Upper right*: lateral incisor (12), first and second premolars (14 & 15) and the second molar (17). *Upper left*: central incisor (21) lateral incisor (22) canine (23) and the second molar (27). *Lower left*: central and lateral incisors (31, 32), canine (33), first premolar (34). *Lower right*: central and lateral incisors (41, 42), canine (43), first and second premolar (44, 45).
iii. **SMB 52B** - subadult - age 10 - 12 years and had 24 permanent and 2 primary teeth – *Upper right*: central and lateral incisors (11, 12), canine (13), first and second premolar - partially erupted (14, 15), first molar (16) and the second molar - partially erupted (17). *Upper left:* central incisor (21), canine - partially erupted (23), first and second premolar - partially erupted (24, 25), first molar (26) and the second molar - partially erupted (27). *Lower left*: central and lateral incisors (31, 32), canine (33), first premolar - partially erupted (34), *primary* second molar (75), first molar (36), second molar - partially erupted (37). *Lower right*: central and lateral incisors (41, 42), canine (43), first premolar - partially erupted (44), *primary* second molar (85), first molar (46) and the second molar - partially erupted (47).
iv. **SMB 4A -** subadult - age range 3.5 to 5.5 years and had 19 primary teeth - *Upper right*: central and lateral incisors (51, 52), canine (53), first and second molar (54, 55). *Upper left*: central and lateral incisors (61, 62), canine (63), first molar (64). *Lower left:* central and lateral incisors (71, 72), canine (73), first and second molar (74, 75). *Lower right:* central and lateral incisors (81, 82), canine (83), first and second molar (84, 85).
v. **SMB 82**: infant - age range 1 to 2 years and had 8 primary teeth – *Upper right*: central and lateral incisors (51 & 52). *Upper left*: central and lateral incisors (61 & 62). *Lower left*: central incisor (71), and the first molar (74). *Lower right*: central incisor (81), and the first molar (84).

The following individual teeth were from the above individuals, which had become loose post-mortem. They were investigated using the SV Micro-CT scanning method.

i. **SMB 73** - 6 permanent teeth - (11, 12, 16, 23, 25, 28),
ii. **SMB 66B** - 6 permanent teeth - (12, 14, 15, 17, 21, 23),
iii. **SMB 52B** - 2 permanent teeth- (11 & 12),
iv. **SMB 4A -** 3 primary teeth- (52, 53, 61),
v. **SMB 82** - 6 primary teeth - (51, 52, 61, 62, 71, 81).

### ETHICS

St Mary’s Anglican Parish requested the excavation of the “free ground” section of the cemetery as they wished to ‘re-use’ the area, and also approved the study of the skeletal remains, Flinders University Social and Behavioural Research Ethics Committee approved the research (SBREC project number 8169). Destructive analysis was not permitted for investigation of the sample (including the dentition and dental arches - alveolar bone tissue) as they are of a rare historical nature.

### METHODS

Large Volume Micro-CT scanning, macroscopic examination, standard (‘plain’) radiography and SV Micro-CT scanning were the modalities compared.

#### a) Large Volume Micro-CT

i. The five skull samples with the dentition in-situ were scanned using the Nikon XT H 225 ST cabinet style micro-CT scanning system (15).
ii. The entire skull (cranium with the mandible in place) of SMB 73 (Fig. 1.) and SMB 52B were scanned. This was undertaken to determine whether scanning the upper and lower jaw opposing each other (i.e., to be visible in the 3D image of the scanned sample), would affect the analysis of the dentition.
iii. The cranium including the maxilla, of adult SMB 66B, subadult SMB 4A, and infant SMB 82, were scanned without the mandible.
iv. The mandible of each individual, SMB 73, SMB 66B (Fig. 2A and 2B.), SMB 52B, SMB 4A and SMB 82, were scanned separately from their associated cranial bones. The mandible being smaller than the entire skull or the cranium alone, allows higher resolution scanning. The spatial resolution (pixel size) selected for each scan was relative to the size of the specimen scanned. Therefore, each complete skull sample (SMB 73, SMB 52B), set of cranial bones with the maxilla (SMB 66B, SMB 4A, SMB 82), or mandible alone (SMB 73, SMB 66B, SMB 52B, SMB 4A, SMB 82), had different scan settings as shown in Table 1

**Figure 1.**
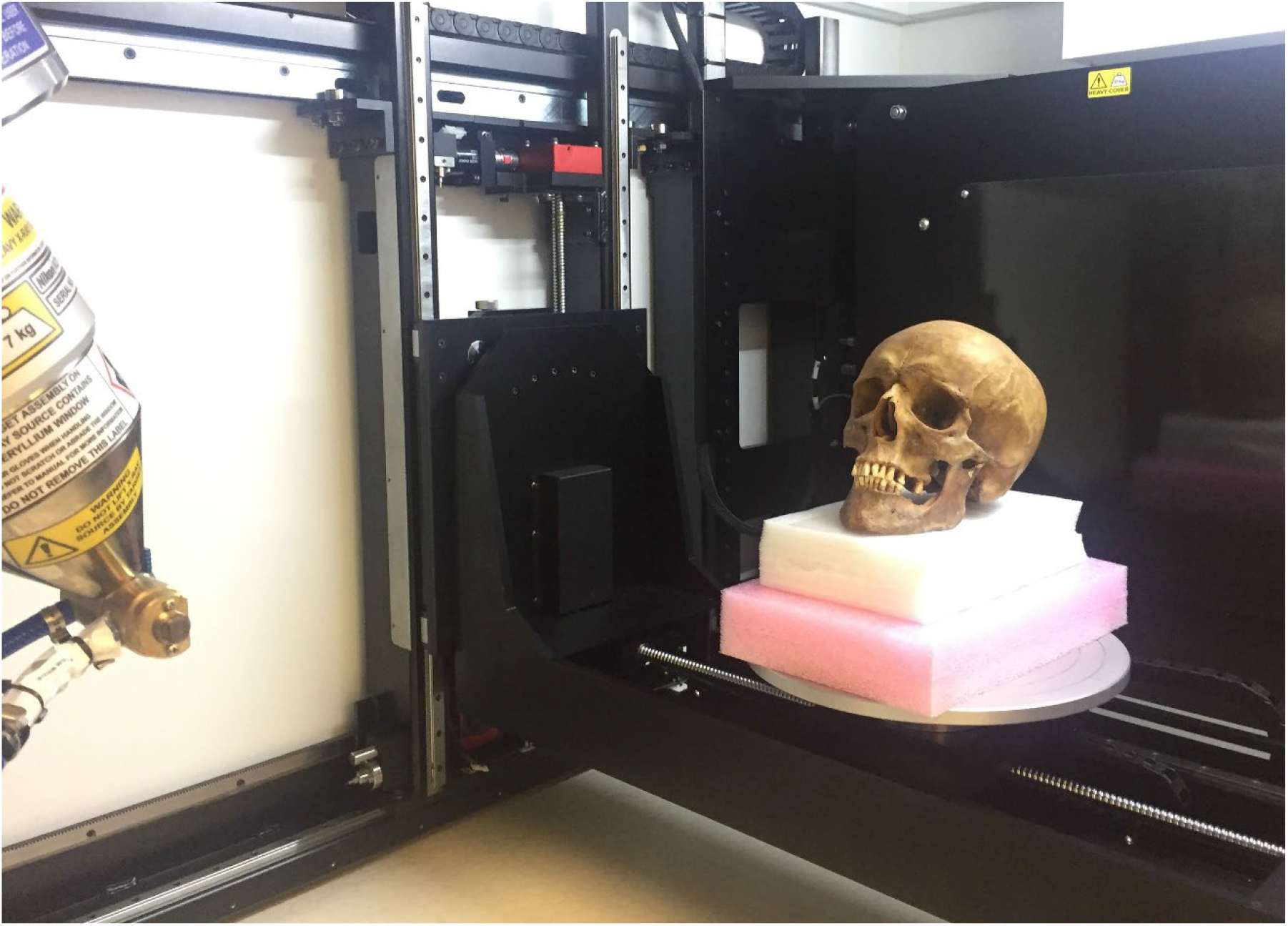
Large Volume Micro-CT. Sample SMB73 - adult male (age 35 - 45 years). Showing the entire skull in position before scanning using the Nikon XH T 225 ST cabinet style system (15).

**Figure 2.**
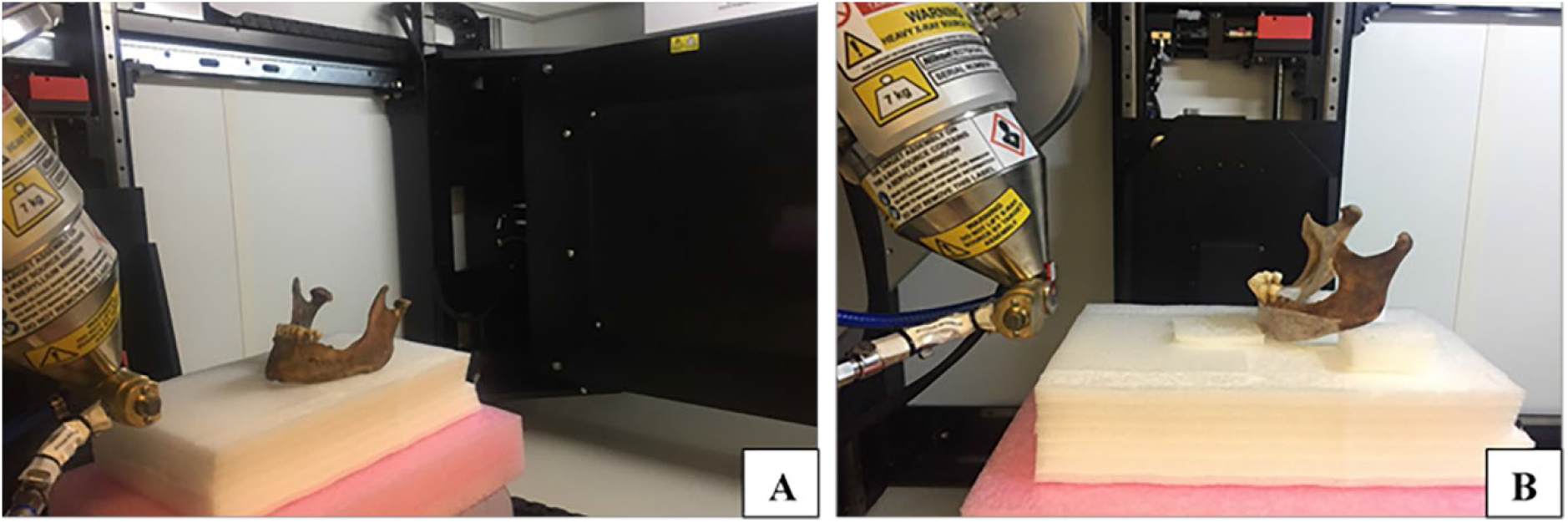
Large Volume Micro-CT. Image **A)** Mandible of SMB 73 - adult male (age 35 - 45 years). Image **B)** Mandible of SMB 66B - adult female (age 35 - 45 years). Showing the position of the in-situ dentition before scanning with Nikon XH T 225 ST cabinet style system (15).

**Table 1.** Large Volume Micro-CT. Settings used for scanning of the in-situ dentition of five archaeological skulls using the Nikon XT H 225 ST Micro-CT scanning system.

| **St Mary's Arch sample ID** | **Skeletal region scanned** | **Pixel size (µm)** | **Source voltage** | **Source current** | **Filter type** |
| --- | --- | --- | --- | --- | --- |
| **SMB 73** | Skull | 60 | 190 | 220 | T |
| **SMB 73** | Mandible | 13 | 190 | 68 | Al |
| **SMB 66B** | Cranium | 50 | 190 | 220 | T |
| **SMB 66B** | Mandible | 13 | 190 | 68 | Al |
| **SMB 52B** | Skull | 60 | 190 | 220 | T |
| **SMB 52B** | Mandible | 22 | 190 | 68 | Al |
| **SMB 4A** | Cranium | 55 | 190 | 220 | T |
| **SMB 4A** | Mandible | 22 | 190 | 68 | Al |
| **SMB 82** | Cranium | 40 | 190 | 220 | T |
| **SMB 82** | Mandible | 18 | 190 | 68 | Al |

**Note:** Skull = cranium and mandible. Cranium = no mandible in place. Mandible = lower jaw only. Filters: Al = Aluminium, T = Tin. Pixel size = spatial resolution

#### b) Processing software - post scanning

The Nrecon software (16), was used for the reconstruction of the data sets produced from the LV Micro-CT scanning process. Avizo 9 (17) data visualisation software was used for image analysis of the reconstructed scan data sets. To increase loading and analysis speed the LV Micro-CT scan data sets were reduced in size (quarter sized data sets were selected). Scoring and/or measures of dental health categories were applied to the 2D and 3D images. The density of teeth and cranium of each of the scanned skulls and mandible was determined by applying a threshold level. This information was stored as an Avizo ‘project’ to provide a consistent and reliable platform for repeated measurements. The digitally reconstructed radiographic (DRR) function (Fig. 3B), together with the ‘clipping’ tool was used to remove a quadrant of the jaw (Fig. 3C), in order to examine all the surfaces of the dentition.

**Figure 3.**
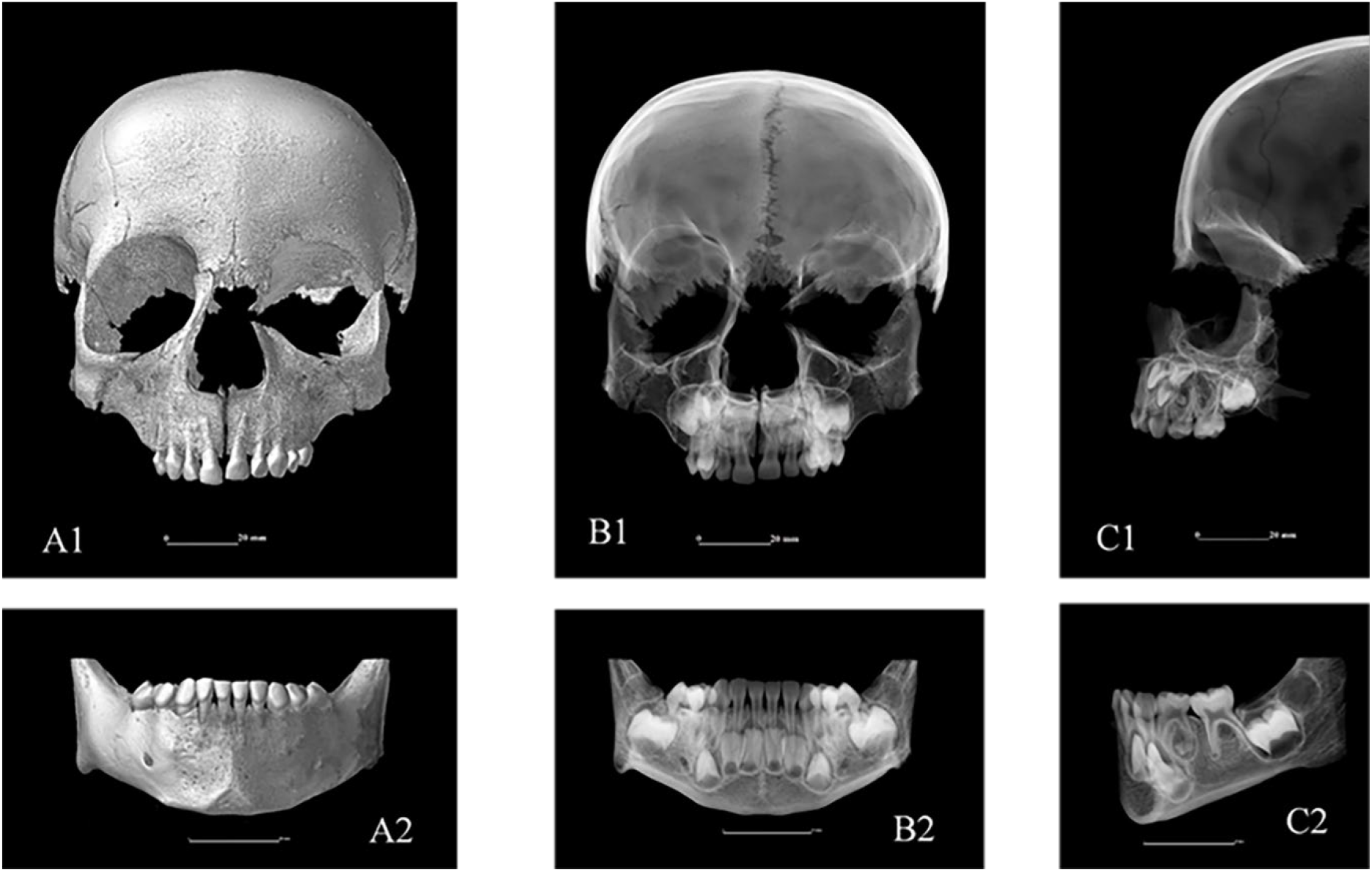
Large Volume Micro-CT. Sample SMB 4A - subadult (3.5 to 5.5 years of age). Images **A1** & **A 2**: Anterior/ labial view of the dentition in-situ in the maxilla and mandible using the Avizo ‘3D’ image function. Images **B1 & B2**: Anterior/ labial view of the same dentition using the Avizo ‘digitally reconstructed radiograph’ (DRR) function (17). Images **C1 & C2:** Lateral view of the dentition using the clipping tool to remove one side of the jaw. Images **B & C** the DRR is showing the developing dentition in the alveolar bones.

### TRADITIONAL METHODS

#### a) Macroscopic examination

i. Visual examination of the dentition and alveolar bone tissues was conducted in a dry laboratory with the aid of a table magnification glass and enhanced lighting.
ii. Enamel opacities were not recorded in this study as enamel hypo-mineralisation developmental defects are difficult to distinguish macroscopically from areas of staining on the tooth caused by the post-mortem environment and/or taphonomic processes (18-20).

#### b) Standard radiographic methods

i. Intraoral periapical and bitewing radiographs were taken using Planmeca X-ray equipment (21), with Phosphor Storage Plates (PSP) as the detectors. Exposure settings were as follows: Tube voltage:70 kV; Tube current: 6mA, with an exposure time of 0.32 seconds.
ii. Extraoral radiographs – the skull of each individual was securely attached to a stand allowing orthopantomogram (OPG) X-ray equipment to rotate around the maxilla and mandibular area. The X-ray source used was a Kavo Pan eXam Plus (22). The tube type: stationary anode; 65kV; Tube current: 15 mA, with an exposure time of up to 16.4 seconds.
iii. Only severe enamel hypoplastic defects can be identified on radiographs and therefore was not assessed using this modality.

#### c) Small Volume Micro-CT

i. Teeth that had fallen out post-mortem or could be removed without any damage to the skull from each individual were investigated. Thus, the same number of teeth and tooth types were *not* available for each individual. Individual tooth specimens were scanned using the SV Micro-CT scanner, the Bruker SkyScan 1276 (23), at 9.0 µm pixel size. The scanner was set at source voltage of 100 kV, source current of 200 µA, and camera binning of 4032 × 2688, with an aluminium and copper filter. The same image processing software as for the LV Micro-CT was used for the reconstruction of the S.V Micro-CT data sets.

### SCORING SYSTEMS AND IDENTIFICATION CRITERIA

Teeth and associated alveolar bones were examined and following observations recorded: i) an inventory of teeth present, ii) tooth wear, iii) dental trauma, iv) class of occlusion, v) caries, including category of radiolucency, vi) alveolar bone status, vii) periodontal disease (horizontal bone loss), viii) enamel hypoplastic defects, ix) interglobular dentine, and x) estimated dental age.

The criteria used for the identification and scoring of defects in teeth and associated alveolar bones of in-situ skulls collected with each investigative method are presented in Table 2.

**Table 2.** Dental and alveolar bone tissue categories. measured for investigation of in-situ dentition, with identification criteria, methods used and systems followed for collection of data, including source for each system

| Categories to be measured | Identification criteria | Method/s to be used | System/s to be followed | Sources |
| --- | --- | --- | --- | --- |
| **Inventory** – total number of teeth in-situ: | Inventory of teeth present | Macroscopic, Radiographs, LV Micro-CT | FDI (ISO 3950) notation system used, data recorded on a visual chart representing the upper and lower permanent/ primary dentition. | ISO, 2009 (24); FDI World Dental Federation, 2022 (25) |
| **Inventory - Ante-mortem** tooth loss | Evidence of healing of alveolar processes | Macroscopic, Radiographs, LV Micro-CT - | Location of healed alveolar where missing tooth type was previously located-recorded (FDI) visual chart used as above | Araújo et al., 2000(26); Kinaston et al., 2019.(27) |
| **Inventory** – Post-mortem tooth loss | No evidence of healing of the alveolar bone - open socket observed | Macroscopic, Radiographs, LV Micro-CT | Location of open socket in alveolar process/ missing tooth type-(FDI) visual chart used as above. | Hillson, 1996 (28) |
| **Occlusion** | Position of jaws & dental arches –in relation to each other Mesio-distally. | Macroscopic, LV Micro-CT | Scored using Angle’s system of classes of malocclusion. | Angle, 1900 p34-44 (29) |
| **Tooth wear** (occlusal) | Evidence of loss of enamel and/or exposure of dentine on the occlusal surface | Macroscopic, SV Micro-CT, LV Micro-CT | Category of tooth wear selected from Molnar’s criteria chart. | Molnar, 1971 (30) |
| **Dental trauma** – | Evidence of morphological damage to the tooth | Macroscopic, SV micro-CT, LV Micro-CT | An adaptation of the index by Winter & Brook was used to record Trauma involving enamel only, enamel & dentine, or enamel, dentine & pulp. | Winter & Brook, 1989 (31) |
| **Caries** | Evidence of decay – on enamel surface, involving enamel & dentine or enamel dentine & the pulp. Identification of a difference in radiolucency | Macroscopic, Radiographs, SV Micro-CT, LV Micro-CT | Tooth type affected (FDI), number and location of caries recorded. Dental probe used for the macroscopic examination. ICDAS/ICCMS categories of radiolucency were selected from a visual chart for radiographic and digitally reconstructed radiographs (DDR) on Avizo 9 software. | Pitts et al., 2013 (14) |
| **Alveolar bone status** | Changes in the structure on the buccal contours of the alveolar margins of the posterior teeth | Macroscopic, LV Micro-CT | Inspection of the buccal contours of the alveolar margins of the posterior teeth. Scored as grade 1 to 4 using Ogden’s (2008) system\. | Ogden, 2008. p.283-307 (32); Riga et al.,2021 (33) |
| **Periodontal disease** | Evidence of horizontal &/or vertical bone loss | Macroscopic, Radiographs, LV Micro-CT | Measurement taken on the midline of the labial/buccal and lingual surfaces of the tooth, from the CEJ to the crest of the alveolar bone. A periodontal probe was used with three-millimetre increments for the macroscopic examination. | Perschbacher, 2014, p 302 (34) |
| **Enamel hypoplastic defects** | Evidence of a enamel hypoplastic defect/s – (linear and/ or pits) | Macroscopic, SV Micro-CT, LV Micro-CT | Using an adaptation of the Enamel Defect Index (EDI), the type of enamel hypoplastic defect/s, the number & location of defects were recorded. A measurement/s of the distance from hypoplastic defect/s to CEJ was taken. | Brook et al., 2001 (35); Elcock et al., 2006.(36) |
| **IGD** (interglobular dentine) | Evidence of areas of defective mineralisation in the dentine structure | SV Micro-CT, LV Micro-CT | Recorded presence of IGD as Yes/No | Colombo et al., 2019 (37); Veselka et al. 2019 (38) |
| **Dental age range estimation** | Eruption rate & position of tooth germs in alveolar bones | Macroscopic, Radiographs, LV Micro-CT | The London Atlas of tooth eruption and development was used to identify the stage of eruption and tooth development. | AlQahtani et al., 2010 (39) ; AlQahtani, 2012 (40) |

Notes: FDI = Fédération Dentaire Internationale. ICCMS = International Caries Classification and Management system. CEJ = Cementum Enamel Junction.

### VALIDITY OF THE SCORING SYSTEM OF CATEGORIES IN TEETH AND ASSOCIATED ALVEOLAR BONES OF IN-SITU SKULLS

The five operators, either post-doctoral staff or post graduate students in Biological Anthropology and/or Archaeology, were trained and independently scored the categories in teeth and associated alveolar bones, using the same methods of investigation. Each operator scored independently, two weeks after the training session. The principal operator (AG) scored the in-situ dentitions, alveolar bone tissues, and individual teeth for the categories (Table 2), macroscopically, on dental radiographs and on the 2D and 3D images produced by the Avizo software from the LV and SV Micro-CT reconstructed scan data sets. All data scoring sessions were repeat tested for the intra-operator reliability two weeks apart. Inter-operator (MM) undertook scoring of the in-situ dentitions, alveolar bone tissues, and individual teeth for the same categories (Table 2), using the comparative methods on different dates and times from intra-operator (AG). Other operators conducted data scoring sessions of the categories listed in Table 2 on images produced by the LV Micro-CT scan data sets only.

### STATISTICAL ANALYSIS

Intra-operator and inter-operator reliability were assessed using Gwet’s Agreement Coefficient (AC1), weighted Gwet’s Agreement Coefficient (AC2) and Intraclass Correlation Coefficient (ICC) using a two-way random-effects model for absolute agreement, for binary and nominal scale data, ordinal scale data and continuous data, respectively.

## RESULTS

### REPRODUCIBILITY - Standard statistical analysis

The agreement between operators (raters) was described and interpreted by running a weighted Gwet’s AC (AC2) analysis. The inter-operator reliability between the methods rating of inventory was moderate to almost perfect agreement, AC2 = 0.90 (95% CI, -0.58 to 1.00), *p* = 0.001, with an observed agreement of 93% (95% CI, 75% to 100%).

The raw data and full results of the standard statistical analysis can be found in the supporting material as:1) S1 Appendix_Gurr et al. 2022_Raw Data_1_EXCEL_Combined Operators_Protected.xlxs spreadsheet, 2) S2 Apprndix_Gurr et al. 2022_Raw Data_2_EXCEL_Combined Operators_Protected.xlxs spreadsheet and 3) S3 Appendix_Reproducibility_Intra_Inter_Operator_Results_.xlsx spreadsheet.

### FINDINGS FOR EACH METHOD TO PROVIDE DATA FOR THE PARAMETERS MEASURED

The ability of each of the investigative methods to provide data required to analyse the dental and alveolar bone tissue health categories is presented in Table 3. The LV Micro-CT scanning method was the *only* single technique that provided information for all of the categories. Macroscopic investigations were limited to the surface structure and could not provide insight on the internal structures of the teeth, thus could not score categories of radiolucency of carious lesions and the presence of IGD (Table 3). Large Volume Micro-CT and macroscopic examination were the only methods that provided relevant information to score the occlusal classification, as the maxillary and mandibular dentitions must be in their natural anatomical position opposing each other. Radiographic and SV Micro-CT techniques did not provide data for at least five of the ten investigated categories (Table 3).

**Table 3.** The ability of each investigative method to provide information on the studied categories of teeth and associated alveolar bones

| Was the method capable of providing data for the following categories?<br>Y/N | Methods |  |  |  |
| --- | --- | --- | --- | --- |
|  | LV Micro-CT | Macroscopic | Radiographic | SV Micro-CT |
| Inventory | Y | Y | Y | N |
| Tooth wear | Y | Y | Y | Y |
| Dental Trauma | Y | Y | Y | Y |
| Occlusion | Y | Y | N | N |
| Caries | Y | Y | Y | Y |
| Alveolar status | Y | Y | N | N |
| Periodontal disease | Y | Y | N | N |
| Enamel hypoplasia | Y | Y | N | Y |
| IGD (dentine defects) | Y | N | N | Y |
| Dental age estimation | Y | Y | Y | N |
| **Technical capabilities:** |  |  |  |  |
| Can the dentition be observed in relation to the skull? | Y | Y | Y | N |
| Is post processing software required? | Y | N | N | Y |
| Are external structures visible? | Y | Y | Y | Y |
| Are internal structures visible? | Y | N | Y | Y |
| 2D image produced? | Y | Y | Y | Y |
| 3D image produced? | Y | Y | N | Y |
| Superimposition issues? | N | N | Y | N |

Note. LV Micro-CT = Large Volume Micro-CT; SV Micro-CT = Small Volume Micro-CT, this method only examined individual tooth samples. IGD = interglobular dentine

### 2) COMPARISON OF THE LEVEL OF SCORING FOR LV MICRO-CT WITH MACROSCOPIC AND RADIOGRAPHIC METHODS

The ‘levels’ of scoring for the information provided by LV Micro-CT, compared to those demonstrated with the macroscopic and radiographic methods for the dental and alveolar bone health categories are summarised in Table 4. All three methods provided identical data for the dental inventory and gave similar results for the estimation of dental age range, for each individual. The score for the radiographic method was consistently at least one category higher for tooth wear compared with the LV Micro-CT and macroscopic examinations. For dental trauma, the macroscopic technique identified the greatest number of episodes (Table 4).

**Table 4.** A comparison of the scoring for LV Micro-CT, with Macroscopic and Radiographic methods of examination of the dentoalveolar complex.

| Categories measured | Methods used |  |  |
| --- | --- | --- | --- |
|  | LV Micro-CT | Macroscopic | Radiographic |
| **Tooth wear**<br>Molnar (1971) (41) grades 1 to 8.<br>min to max range observed: | 1 to 5 | 1 to 5 | 2 to 5 |
| **Dental trauma**<br>Winter & Brook, (1989) (31)<br>total number of teeth affected: | 9 | 17 | 10 |
| **Class of Occlusion**<br>Angle, (1898) (29)<br>range of classes 1 to 3 | Class 1 | Class 1 | N/A |
| **Caries**<br>*total number of lesions observed | 56 | 80 | 41 |
| **Caries**<br>ICDAS/ICMS (14) radiolucency<br>categories 0 to 6 min to max range<br>observed | 1 to 4 | N/A | 1 to 4 |
| **Enamel hypoplastic defects**<br>Brook et al., (2001) (35)<br>total number of teeth affected | 44 | 13 | N/A |
| **IGD** (interglobular dentine)<br>Colombo et al., (2019) (37);<br>Veselka et al., (2019) (38),<br>total number of teeth affected | 7 | N/A | N/A |

**Notes.** \*More than one carious lesion may be present on a tooth. IGD = Interglobular dentine. Raw data giving further details in each category are available in the supporting material as: 1) S1 Appendix\_Gurr et al. 2022\_Raw Data\_1\_EXCEL\_Combined Operators\_Protected.xlsx spreadsheet, and 2) S2 Appendix\_Gurr et al. 2022\_Raw Data\_2\_EXCEL\_Combined Operators\_Protected.xlsx spreadsheet

Alveolar bone status grades were higher for the LV Micro-CT method compared with the macroscopic examination results. This category could not be scored on radiographs. The measurements from the Cemto-enamel Junction (CEJ) to the crest of the alveolar bone, for horizontal bone loss as indicating periodontal disease, provided by the LV Micro-CT were longer than those obtained using the macroscopic method. The ranges of measurements for horizontal bone loss in sample SMB 73, were 2.04 to 7.36 mm, with the LV Micro-CT method, compared with the 2 to 6 mm by the macroscopic method.

A higher total number of carious lesions was identified by the macroscopic method (Table 4). The scoring of the category of radiolucency of carious lesions (14), for the LV Micro-CT and radiographic methods was identical (Table 4); this category could not be scored macroscopically. More individuals were identified with evidence of enamel hypoplastic (EH) defects by the LV Micro-CT method (Fig. 4.), compared with the macroscopic examination (Table 4). The maximum distance measurement for the location of an EH defect from the CEJ provided by the LV Micro-CT method was greater than that obtained by the macroscopic examination. Only the LV Micro-CT technique provided images of IGD that could be scored.

**Figure 4.**
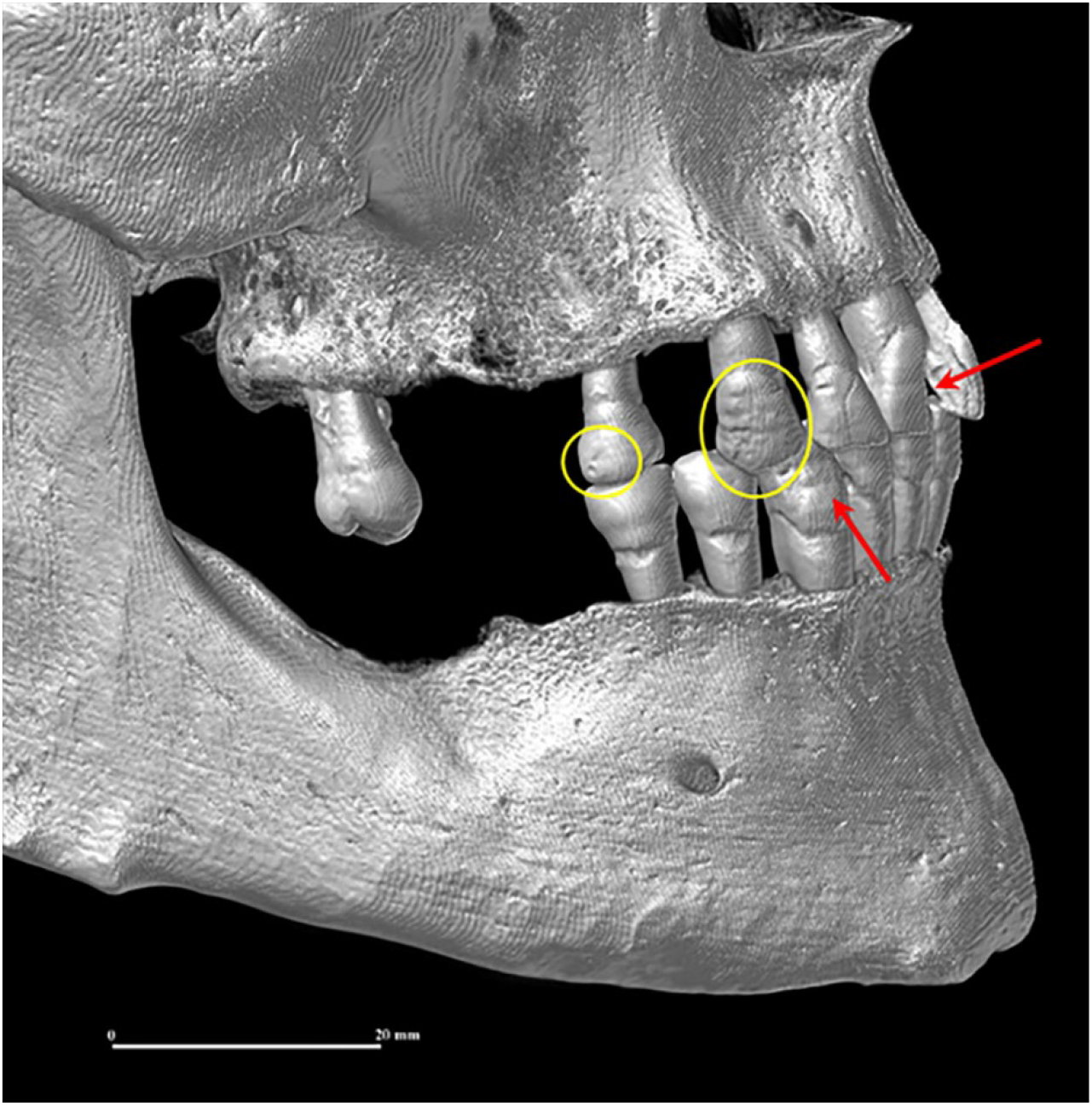
Large Volume Micro-CT. Sample SMB 73, adult male (age 35-45 years) **Image**: Lateral view of the dentition in-situ in the maxilla and mandible, showing multiple examples of enamel hypoplastic defects (linear = *red arrows* and pits = *yellow circles*). Using Avizo 9 post processing software.

### 3) COMPARISON OF THE LEVEL OF SCORING FOR LV MICRO-CT WITH THE SV MICRO-CT METHOD

The SV Micro-CT method requires individual teeth that are separated from the jaw and could not provide information for scoring of: i) inventory, ii) class of occlusion, iii) alveolar bone status, iv) periodontal disease and v) estimation of dental age (Table 3). A summary of scoring of the information provided by the LV Micro-CT compared with that from the SV Micro-CT scanning system for the remaining categories is presented in Table 5.

**Table 5.** A comparison of scoring for the LV Micro-CT with the SV Micro-CT scanning methods of individual tooth samples.

| Categories | Methods |  |
| --- | --- | --- |
|  | LV Micro-CT | SV Micro-CT |
| **Tooth wear**<br>Molnar (1971) (41) grades: 1 to 8.<br>min to max range observed: | 1 to 4 | 1 to 5 |
| **Dental trauma**<br>Winter & Brook, (1989) (31)<br>total number of teeth affected: | 2 | 17 |
| **Caries**<br>*total number of lesions observed | 17 | 15 |
| **Caries**<br>ICDAS/ICMS (14) radiolucency categories 0 to 6<br>min to max range observed | 1 to 3 | 1 to 3 |
| **Enamel hypoplastic defects**<br>Brook et al., (2001) (42)<br>total number of teeth affected | 28 | 45 |
| **IGD** (interglobular dentine)<br>Colombo et al., (2019) (37); Veselka et al.,<br>(2019) (38)<br>total number of teeth affected | 0 | 6 |

**Note.** \*More than one carious lesion may be present on a tooth. Raw data giving further details in each category are available in the supplementary material as:
1) S1 Appendix\_Gurr et al. 2022\_RAW DATA\_1\_EXCEL\_COMBINED OPERATORS\_PROTECTED.xlsx spreadsheet and 2) S2 Appendix\_Gurr et al. 2022\_RAW DATA\_2\_EXCEL\_COMBINED OPERATORS\_PROTECTED.xlsx spreadsheet.

Large Volume Micro-CT and SV Micro-CT methods produced similar or identical results for several of the categories. For example, the ICCMS range of scores for the category of radiolucency of carious lesions was the same for both methods (i.e., 1 to 3) (Table 5), and the total number of carious lesions identified by the LV Micro-CT method was 17 lesions, while that for the SV Micro-CT system was 15 lesions (Table 5). However, more incidences of dental trauma, EH and IGD (Fig. 5.) were identified by the SV Micro-CT scanning method compared with the LV Micro-CT (Table 5) system.

**Figure 5.**
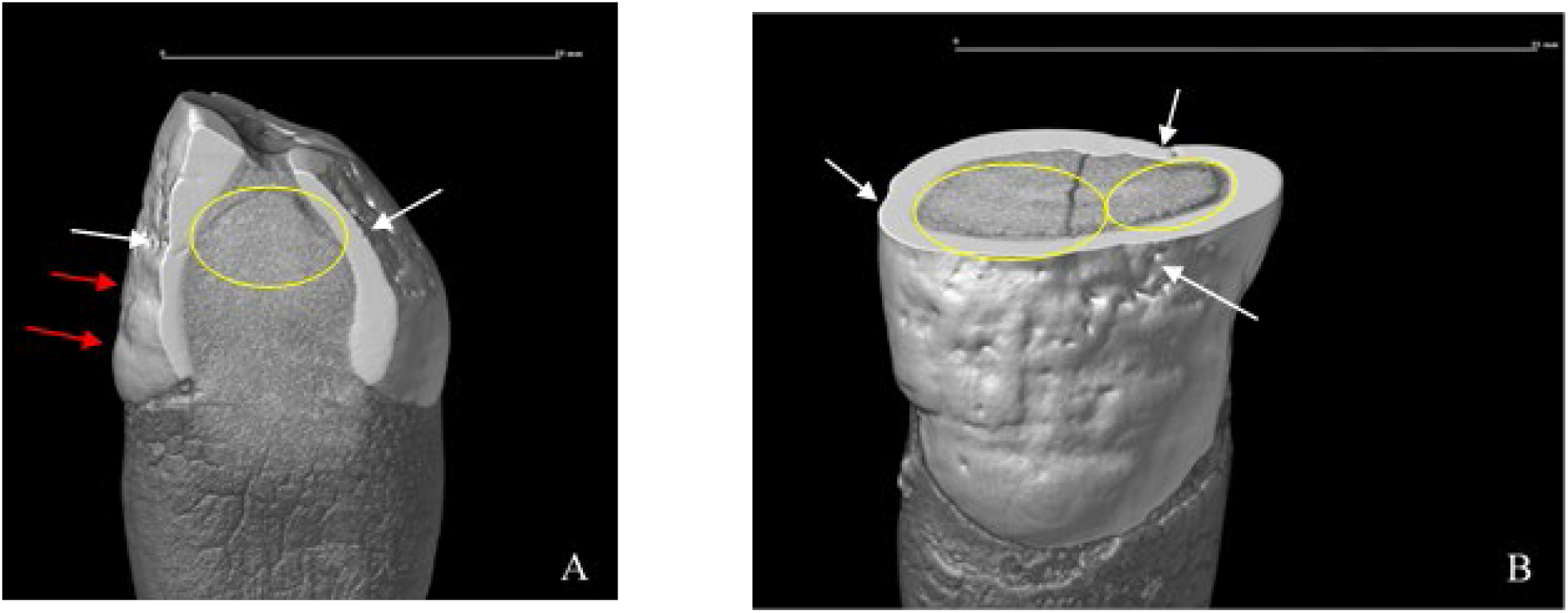
Small Volume Micro-CT. Sample SMB 73 - adult male (age 35-45 years). Individual tooth (upper left canine- FDI - 23). Image **A**. Sagittal view (slice) of the external and internal structures of the tooth. This tooth has enamel hypoplastic defects (EH) pits – *white arrows*, and linear enamel hypoplastic defects - r*ed arrows*, and areas of interglobular dentine (IGD) -*yellow circle*. The areas of IGD had occurred at a similar level to the EH on the crown of this tooth. Image **B**. Transverse view (slice) of the crown. EH and IGD, which had occurred at the same level of the tooth structure. Data visualisation was using Avizo 9 software.

## DISCUSSION

Our investigations indicate that while each method, LV Micro-CT, macroscopic, radiographic and SV Micro-CT, has its applications, there are differences in the categories that they can score and in the details that they can provide. In this discussion the methods are considered in relation to the order of the aims of the study.

### 1) Large Volume Micro-CT

The innovative non-destructive LV Micro-CT scanning system is a valuable technique for the analysis of the dentoalveolar complex in archaeological samples. It provided overall information for the full range of categories measured, (Tables 3, 4, 5), fulfilling the first aim of this study. The volumetric capacity of the LV Micro-CT allowed it to accommodate a whole skull (Fig.1), with dentition in-situ and generated adequate information to investigate in detail: tooth wear, dental trauma, classification of occlusion, caries, alveolar bone status, periodontal disease, enamel hypoplasia, interglobular dentine and dental age estimation (Tables 3, 4 and 5; Fig. 3, 4 and 6). The post scan processing software (Avizo 9) (17), necessary to analyse the LV Micro-CT scan data sets, was chosen for this study for the multiple features that it offered such as the ‘Digital Reconstructed Radiograph’ (DRR) function and its ability to examine the external and internal structures of the dentoalveolar complex simultaneously in 3D. The abilities of the computer software enhanced the detailed investigation of the above categories in teeth, the associated alveolar bones and developmental stages of un-erupted teeth using the LV Micro-CT system (Tables 4 and 5; Fig. 3, 4 and 6). Scoring of the LV Micro-CT scan images using the Avizo software can be undertaken with training by operators of a non-dental background.

**Figure 6.**
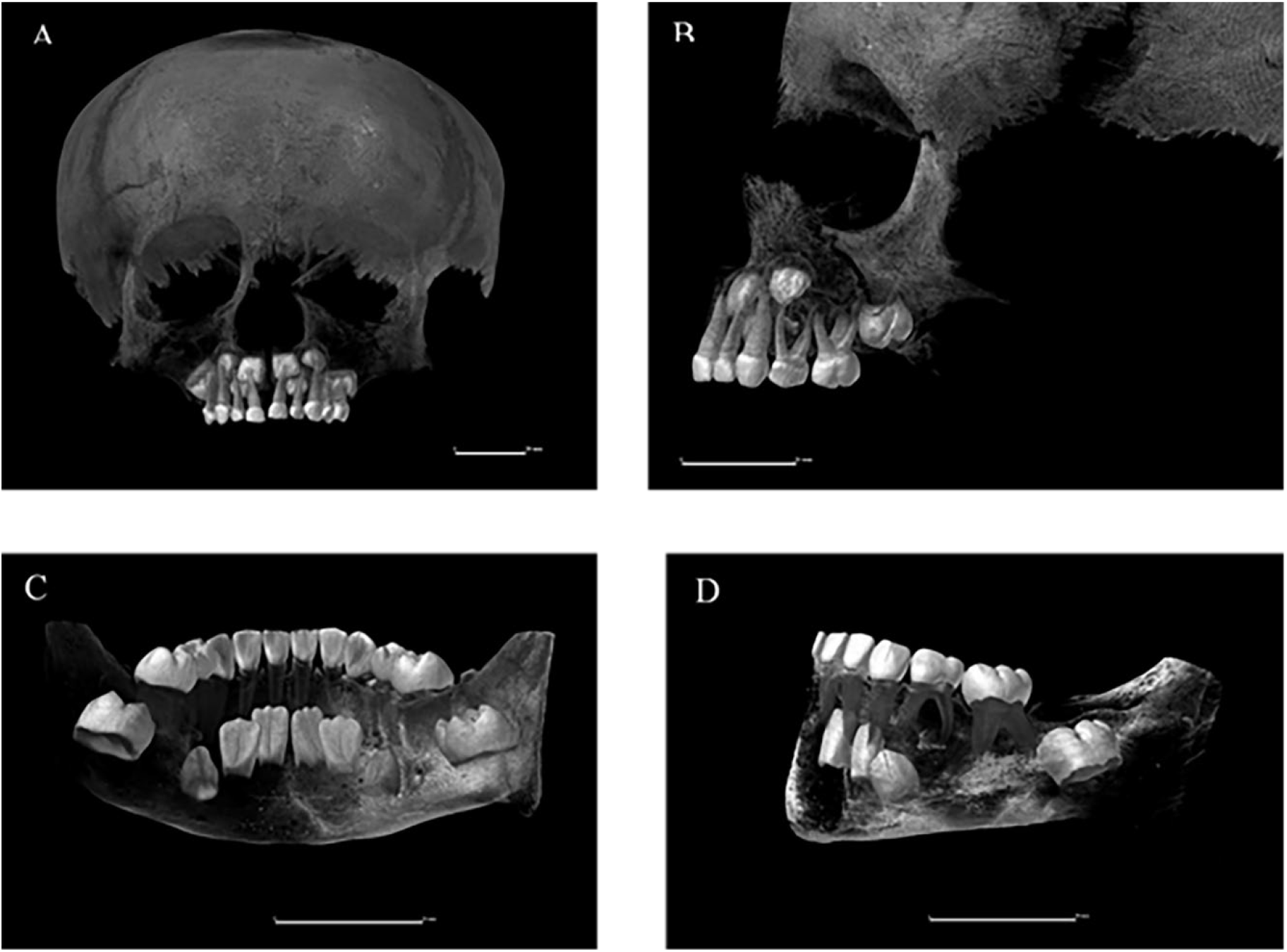
Large Volume Micro-CT. Sample SMB 4A (age 3.5 - 4.5 years). Image **A**: Anterior/ labial view of the dentition in-situ in the maxilla. Manipulation of the bone density threshold level showed the developing tooth germs in the alveolar bone. **Image B**. Lateral view of the same dentition. Image **C**: Posterior/ lingual view of the mandible showing erupted and unerupted teeth developing in the jaw. Image **D** Lateral view of the mandible showing erupted and unerupted teeth developing in the jaw.

Information, that is additional to the categories studied here, can be gained from the LV Micro-CT scan of a skull which can provide an insight into the changes that occur in the dentition and the possible association of bony changes (developmental and pathological conditions), that can affect both the teeth and alveolar bones. This is important as teeth and dental arches are separate complex systems of two or more structures (43-45). To determine the value of the LV Micro-CT technique for the analysis of dentitions at different stages of development, the wide age range of the individuals was deliberately chosen. An example of developmental changes seen in the teeth of the adults and subadults was a marked contrast in the density threshold level of the dentine and the enamel for the scoring of one parameter (i.e., IGD). This was due to the differing stages of mineralisation of their dentitions (46). The LV Micro-CT scanning method also provides data for an overall impression of health through the examination of the cranium and the identification of co-morbidities such as pathological manifestations in the bone, for example cribra orbitalia (47-49), or caries sicca (50). Dentition developing during an extended health insult could be affected (27, 51-62).

### 2) Comparison of Large Volume Micro-CT, Macroscopic and Radiographic methods

Differences and similarities were seen in the ‘level ‘of scoring between the LV Micro-CT, macroscopic and radiographic methods. All three methods produced identical scores for the inventory of the archaeological dentitions. This was to be expected as this category is not open to interpretation. The macroscopic method examined the external 3D structures of the dentition in-situ in the skull. This method identified a higher number of traumatic injuries to the dentitions and carious lesions (Table 4), compared with the other methods. The disadvantage of this method is its inability to examine internal structures, as only normal light is used to view the real specimen. The other methods use X-rays to generate images of the real specimen, including those of the internal structures (Fig. 5). Therefore, confirmation or scoring of any changes in the density of the internal structures of the teeth associated with a carious lesion cannot be undertaken with the macroscopic method.

Tooth wear grades were consistently higher on radiographs compared with the other methods. This may have been due to issues associated with superimposition (i.e., one structure being superimposed on top of another), on the 2D radiographs. Distortion of the radiographic image can occur, especially in the premolar and molar cusps regions that could have influenced the scores. To overcome superimposition during the image analysis of the LV Micro-CT 2D digitally reconstructed radiographs (DRR), using the Avizo 9 software (Fig. 3B and 3C), the ‘clipping’ tool was used to remove a quadrant of the jaw or individual teeth. The location of caries could be determined by manipulating the DRR images on the computer screen, which allowed all surfaces of the tooth to be examined. The use of these software tools, however, did not influence the range of scores given for the ICCMS (International Caries Classification and Management system) (14), category of radiolucency for caries, as they were similar for both the LV Micro-CT and Radiographic methods (Table 4).

Enamel hypoplastic (EH) defects that were not seen during the macroscopic examination of the teeth of one subadult (SMB 52B), were identified by the LV Micro-CT scanning method. Furthermore, the LV Micro-CT method identified a higher number of EH defects in total for the five individuals compared with the macroscopic analysis (Table 4). This increase in the identification of EH was likely due to the high-resolution 3D images produced by the LV Micro-CT and the ‘zoom’ function of the software. The resolution of the scan images could have affected the measurements taken for the location of EH defects (i.e., distance from the CEJ) as these were consistently higher compared with the measurements taken for the macroscopic examination. Radiologically, EH is difficult to identify except in the most severe cases.

A similar increase in the length of the measurements taken using the LV Micro-CT method for evidence of horizontal alveolar bone loss (suggestive of periodontal disease) (Table 3), compared with the macroscopic method, was seen. For example, there was a difference of 1.36 mm between the LV Micro-CT and the macroscopic measurements for one adult individual (SMB 73). While this difference is not large the ability of the measurement function on the Avizo software to present data at as many points past the decimal as required, means the measurements taken are more accurate than those taken using the periodontal probe. Once more, the higher-resolution of the LV Micro-CT images assisted in the identification of the precise location of the CEJ and the crest of the alveolar bone.

The three compared methods all provided similar dental age ranges for the five individuals. The 2D DRR images (Fig. 3B and 3C), used with the LV Micro-CT method, were a direct comparison with the radiographic method for this parameter. The DRR images from the LV Micro-CT scanning data sets were of high resolution (Figure 3) and of superior quality to the radiographic method. The 3D image analysis of the LV Micro-CT data sets using Avizo software could reduce the bone density threshold of the maxilla and mandible to reveal denser structures beneath, such as the developing permanent tooth germs in the jaw (Fig. 6). Changing the bone density threshold on the 3D images can provide information on the position and stage of development of primary and/or permanent teeth in the jaws. The macroscopic method can only provide an estimation of dental age on the evidence of the erupted or erupting dentition.

Addressing the second aim, the LV Micro-CT was the *only* technique to provide data required to complete the analysis of the dentoalveolar complex for the five individuals (Table 3). The macroscopic method could not examine the internal structures of teeth and therefore was not able to provide all data required (Table 3 and 4), and the radiographic method provided the least amount of data (Table 3 and 4). Combined, the macroscopic and radiographic methods can provide data for the majority of the parameters except for IGD (Table 3). However, the collection and analysis of data from two methods is time consuming and for several of the parameters, such as Occlusal classification, Alveolar status, Periodontal disease, and EH, only the macroscopic method can provide measurements. Thus, only the LV Micro-CT scanning system, using the appropriate post processing software, can provide data for all the categories measured.

### 3) Comparison of ‘level’ of scoring between Large Volume Micro-CT and Small Volume Micro-CT scanning methods for individual loose teeth

Findings of the individual teeth, not in the alveolar bone, showed similarities and differences in the ‘level;’ of scoring between the LV Micro-CT and SV Micro-CT methods. Scoring for tooth wear, total number of carious lesions and categories of radiolucency for carious lesions were similar or the same for these two methods (Table 5). However, a higher number of episodes of dental trauma, EH defects and areas of IGD were identified using the SV Micro-CT scanning system (Table 5). This included evidence of dental trauma and EH defects that had not been observed using the LV Micro-CT method for subadult SMB 4A.

The placement of the jaws and the close proximity of the occlusal surfaces of the anterior teeth (FDI - 11, 12, 13, 21, 22, 23), of SMB 73, during LV Micro-CT scanning process (Fig. 1), did affect the image analysis of these data sets. The examination of the dentine for areas of IGD requires the removal of slices of the scanned specimen to analyse the structures below. The ability to identify if areas of IGD were present in the region of the dentine closest to the tip of the crown was more difficult than for the examination of the dentition in the mandible that was scanned separately from the maxilla (Fig. 2). The areas of IGD that were identified using the LV Micro-CT method were all in the mandibular dentition. The SV Micro-CT method identified areas of IGD in 5 of the 6 maxillary tooth samples for sample SMB 73 (Fig. 5), compared with the LV Micro-CT method. It is probable that some findings from the SV Micro-CT scanning system (Table 3), were due to the ‘higher’ resolution achievable compared with the LV Micro-CT system and the fact that no other teeth are in close proximity to the specimen under investigation.

Fulfilling aim 3, the LV Micro-CT provides data for all categories measured in this study, but if ‘higher’ resolution analysis of specific conditions such as EH and/or IGD on individual tooth samples that are no longer present in the jaw (Fig. 5), is required, the SV Micro-CT method can provide more detailed data. This is due to the size of the specimen under investigation as it is directly related to the achievable resolution of the SV Micro-CT scanning system (63, 64).

### Advantages and Limitations of the Large Volume Micro-CT scanning method

The LV Micro-CT method can be used as a stand-alone technique for the investigation of dental and alveolar bone tissues. This method preserves rare and/or delicate archaeological skull samples, as well as other associated bony specimens, for future research. The scanned data set of a specimen can be digitally archived via an international database. Alternatively, the data sets can be converted to STL files for 3D printing.

There are many types of software available for use with LV Micro-CT scan data sets and the choice will undoubtedly increase in the future. Some of the current options could be used for triaging scan images. For example, Dataviewer (23), (a free downloadable software), is ideal as a screening tool as it provides slice by slice images in grayscale. Compared with the Avizo software Dataviewer is somewhat simplistic, however, far less processing power is required to achieve results. Other software commonly used for the analysis of the L.V Micro-CT scan data sets are CTvox (65), CTvol (65) and CTan (65).

The costs associated with the LV Micro-CT scanning method, the availability of the equipment and the requirement for an expert licensed operator to conduct the scanning process currently limit its use. However, if precise analysis of a specific category is not the focus, the LV Micro-CT system could be more economical in terms of the cost than SV Micro-CT for the full dentoalveolar complex as it is able to scan a whole skull. The large data sets produced require substantial computer power for analysis but reducing the scan data sets will allow a more rapid analysis, though it will decrease the resolution.

### Limitations of this study

The small sample size meant that many of the results for the statistical analysis (Gwet’s AC & AC2) had a confidence interval of 1.00, which indicates a perfect agreement by the operators (raters). When there is not perfect agreement most results have very wide confidence intervals. This is again due to the very small sample size. Where a result is presented without a confidence interval, e.g. 0.75 (.,.) it indicates that there were not enough results to calculate the confidence interval. There was not enough variation in the data to provide results for occlusion, and there was not enough variation in IGD. Not all statistics could be estimated due to not enough data or not enough variation in the data. No variation, however, is expected to occur when there is a perfect agreement of ratings.

## CONCLUSION

The LV Micro-CT technique is an advance from the current non-destructive methods for the study of the dentition and the surrounding tissues in archaeological samples. Macroscopic and radiographic methods alone could not identify all the categories but were able to identify the majority when these two methods were combined. The SV Micro-CT provided higher resolution analysis for loose individual teeth due to the small size of the specimens. The SV Micro-CT technique could not study the supporting alveolar bone tissues.

## Data Availability

All data produced in the present study are available upon reasonable request to the authors.

## ACKNOWLEDGEMENTS

The authors thank Fr. William Deng, of St Mary’s Anglican Church for continued access to the St Mary’s skeletal sample. The authors acknowledge the facilities and the scientific and technical assistance, of the Australian Microscopy & Microanalysis Research Facility at the Adelaide Microscopy, University of Adelaide. Dr Agatha Labrinidis for assistance with the Bruker SkyScan 1276 Micro-CT scanning system. Prof Egon Perilli, from the Biomedical Engineering Medical Device Research Institute, Flinders University, for the knowledge he kindly shared regarding Large Volume Micro-CT systems. Thank you to Flinders Microscopy and Microanalysis (FMMA), for access to the brand-new Nikon L.V Micro-CT scanning system. The authors thank Dr Derek Lerche from the Adelaide Dental School for his assistance with the dental radiographs. Kelly Hall, from Adelaide Health Technology Assessment, School of Public Health, The University of Adelaide, provided assistances with statistical analysis of the study data. We are grateful for the assistance of Dr John Wetherell, Claudia Barrientos and Ella Kelty with the reproducibility scoring.

## Notes

### Competing Interest Statement

The authors have declared no competing interest.

### Funding Statement

This study was funded by the University of Adelaide, Adelaide Dental School Research Committee South Australia

### Author Declarations

ETHICS St Marys Church Anglican Parish South Australia, requested the excavation of the 19th century free ground section of the cemetery as they wished to re-use the area and also approved the study of the skeletal remains Flinders University Social and Behavioural Research Ethics Committee approved the research (SBREC project number 8169). Destructive analysis was not permitted for investigation of the sample (including the dentition and dental arches - alveolar bone tissue) as they are of a rare historical nature. The excavated site within the cemetery was unmarked therefore the excavated individuals could not be identified.

### Summary of Updates

The order of the authors.

## REFERENCES

1. Gurr A, Brook AH, Kumaratilake J, Anson T, Pate FD, Henneberg M. Was it worth migrating to the new British industrial colony of South Australia? Evidence from skeletal pathologies and historic records of a sample of 19th-century settlers. International journal of paleopathology. 2022;37:41–52.

2. Gurr A, Kumaratilake J, Brook AH, Ioannou S, Pate FD, Henneberg M. Health effects of European colonization: An investigation of skeletal remains from 19th to early 20th century migrant settlers in South Australia. PloS one. 2022;17(4):e0265878–e.

3. Kramer B, Molema K, Hutchinson EF. An osteological assessment of cyclopia by micro-CT scanning. Surgical and radiologic anatomy (English ed). 2019;41(9):1053–63.

4. Main M, Tan J, Yong R, Williams R, Labrinidis A, Anderson PJ, et al. Craniofacial Phenomics: Three-Dimensional Assessment of the Size and Shape of Cranila and Dentofacial Structures. In: Dworkin S, editor. Craniofacial Development Methods in Molecular Biology. 2403. New York: Humana; 2021.

5. Tan J, Labrinidis A, Williams R, Main M, Anderson PJ, Ranjitkar S. Micro-CT-Based Bone Microarchitecture Analysis of the Murine Skull. In: Dworkin S, editor. Craniofacial Development Methods of Molecular Biology. 2403. New York: Humana; 2022.

6. Welsh H, Nelson AJ, van der Merwe AE, de Boer HH, Brickley MB. An Investigation of Micro-CT Analysis of Bone as a New Diagnostic Method for Paleopathological Cases of Osteomalacia. International journal of paleopathology. 2020;31:23–33.

7. Alsop K, Norman DG, Baier W, Colclough J, Williams MA. Advantages of micro-CT in the case of a complex dismemberment. Journal of forensic sciences. 2022.

8. Rutty GN, Brough A, Biggs MJP, Robinson C, Lawes SDA, Hainsworth SV. The role of micro-computed tomography in forensic investigations. Forensic science international. 2012;225(1):60–6.

9. Appleby JD, Rutty GNP, Hainsworth SVP, Woosnam-Savage RCBA, Morgan BP, Brough AP, et al. Perimortem trauma in King Richard III: a skeletal analysis. The Lancet (British edition). 2015;385(9964):253–9.

10. Fraberger S, Dockner M, Winter E, Pretterklieber M, Weber GW, Teschler-Nicola M, et al. Micro-CT evaluation of historical human skulls presenting signs of syphilitic infection. Wiener Klinische Wochenschrift. 2021;133(11-12):602–9.

11. Nikolova S, Toneva D, Georgiev I, Lazarov N. Sagittal suture maturation: Morphological reorganization, relation to aging, and reliability as an age-at-death indicator. American Journal of Physical Anthropology. 2019;169(1):78–92.

12. Booth TJ, Redfern RC, Gowland RL. Immaculate conceptions: Micro-CT analysis of diagenesis in Romano-British infant skeletons. Journal of archaeological science. 2016;74:124–34.

13. Morgan JA. The Methological and Diagnostic Applications of Micro-CT to Paleopathology: A Quantative Study of Porotic Hyperostosis [Electronic]. Ontario, Canada: The University of Western Ontario; 2014.

14. Pitts NB, Ekstrand KR. International Caries Detection and Assessment System (ICDAS) and its International Caries Classification and Management System (ICCMS) - methods for staging of the caries process and enabling dentists to manage caries. Community dentistry and oral epidemiology. 2013;41(1):e41–e52.

15. Nikon Metrology. Products: X-ray and CT Technology: X-ray and CT systems: 225kV and 320kV CT inspection and metrology. 2021 [Available from: https://www.nikonmetrology.com/de/roentgen-und-ct-technik/x-ray-ct-syteme-225kv-und-320kv-ct-inspektion-und-messtechnik.

16. Micro Photonics Inc. Micro-CT Software: NRecon Reconstruction Software: Fast micro-CT reconstruction software.: Micro Photonics Inc.; 2020 [Available from: https://www.microphotonics.com/micro-ct-systems/nrecon-reconstruction-software/.

17. ThermoFisher Scientific. 3D Visulaization & Analysis Software: Amiro-Avizo 2019 [Available from: https://www.thermofisher.com/au/en/home/industrial/electron-microscopy/electron-microscopy-instruments-workflow-solutions/3d-visualization-analysis-software.html.

18. Efremov JA. Taphonomy: New Branch of Paleontology. American Geologist. 1940;74(2):81–93.

19. Garot E, Couture-Veschambre C, Manton D, Beauval C, Rouas P. Analytical evidence of enamel hypomineralisation on permanent and primary molars amongst past populations. Scientific reports. 2017;7(1):1712–10.

20. Garot E, Couture-Veschambre C, Manton D, Bekvalac J, Rouas P. Differential diagnoses of enamel hypomineralisation in an archaeological context: A postmedieval skeletal collection reassessment. International journal of osteoarchaeology. 2019;29(5):747–59.

21. PLANMECA. Intraoral X-ray unit: Planmeca OY; 2022. https://www.planmeca.com/imaging/intraoral-imaging/intraoral-x-ray-unit/.

22. KaVo Dental GmbH. KaVo Imaging Solutions: KaVo Pan eXam Plus.: KaVo Dental GmbH; nd. https://www.henryschein.be/be-nl/images/dental/Pan_eXam_Plus_ENG.pdf.

23. Bruker. Products: http://bruker-microct.com/products/downloads.htm. 2018. http://bruker-microct.com/products/downloads.htm.

24. International Organisation for Standardization, American National Standard, American Dental Association. Specification No. 3950. Designation System for Teeth and Areas of the Oral Cavity Switzerland: American Dental Association; 2010. https://webstore.ansi.org/preview-pages/ADA/preview_ISO+ANSI+ADA+Specification+No.+3950-2010.pdf.

25. FDI World dental Federation. FDI Two-Digit Notation France: FDI World Dental Federation; 2022. : http://www.fdiworldental.org/resources/5_0notation.html.

26. Araújo MG, Silva CO, Misawa M, Sukekava F. Alveolar socket healing: what can we learn? Periodontology 2000. 2015;68(1):122–34.

27. Kinaston R, Willis A, Miszkiewicz JJ, Tromp M, Oxenham MF. The Dentition: Development, Disturbances, Disease, Diet, and Chemistry. In: Buikstra JE, editor. Ortner’s Identification of Paleopathological Conditions in Human Skeletal Remains. Third ed. London: Academic Press; 2019.

28. Hillson S. Dental Anthropology. Cambridge: Cambridge University Press; 1996.

29. Angle EH. Treatment of malocclusion of the teeth and fractures of the maxillae: Angle’s system. 6th ed., greatly enl. and entirely rewritten, with 299 ill. ed. Philadelphia: S. S. White Dental Mfg. Co; 1900.

30. Molnar S. Human tooth wear, tooth function and cultural variability. American Journal of Physical Anthropology. 1971;34(2):175–89.

31. Winter GB, Brook AH. Injuries (Traumactic). In: Rowe AHR, Alexander AG, Johns RB, editors. A Comprehensive Guide to Clinical Dentistry. A Companion to Dental Studies. London: Class Publishing; 1989. p. 104–11.

32. Ogden AR, Pinhasi R, White WJ. Gross enamel hypoplasia in molars from subadults in a 16th-18th century London graveyard. American journal of physical anthropology. 2007;133(3):957–66.

33. Riga A, Begni C, Sala S, Erriu S, Gori S, Moggi-Cecchi J, et al. Is root exposure a good marker of periodontal disease?. Bulletin of the International Association of Paleodontology. 2021;15(1):21–30.

34. Perschbacher S. Periodontal Diseases. In: White SC, Pharoah MJ, editors. Oral Radiology: Prinicplas and Interpretations. St Louis: Elsvier; 2014. p. 299–313.

35. Brook AH, Elcock C, Hollonsten A-L, Poulsen S, Andreasen J, Koch G, et al. The development of a new index to measure enamel defects. 12th International Symposium on Dental Morphology 2001 2001. p. 59–65.

36. Elcock C, Lath DL, Luty JD, Gallagher MG, Abdellatif A, Backman B, et al. The new enamel defects index: testing and expansion. Eur J Oral Sci. 2006;114:35–8.

37. Colombo A, Ortenzio L, Bertrand B, Coqueugniot H, KnüSel CJ, Kahlon B, et al. Micro-computed tomography of teeth as an alternative way to detect and analyse vitamin D deficiency. Journal of Archaeological Science: Reports. 2019;23:390–5.

38. Veselka B, Brickley MB, D’Ortenzio L, Kahlon B, Hoogland MLP, Waters-Rist AL. Micro-CT assessment of dental mineralization defects indicative of vitamin D deficiency in two 17th-19th century Dutch communities. Am J Phys Anthropol. 2019:1–10.

39. AlQahtani SJ, Hector MP, Liversidge HM. Brief communication: The London atlas of human tooth development and eruption. American Journal of Physical Anthropology. 2010;142(3):481–90.

40. AlQahtani S. The london atlas: developing an atlas of tooth development and testing its quality and performance measures. ProQuest Dissertations Publishing; 2012.

41. Molnar S, Przybeck TR, Gantt DG, Elizondo RS, Wilkerson JE. Dentin apposition rates as markers of primate growth. American Journal of Physical Anthropology. 1981;55(4):443–53.

42. Brook AH, Elcock C, Hallonsten AL, Poulsen S, Andreasen J, Koch G, et al. The development of a new index to measure enamel defects. In: Brook AH, editor. Dental Morphology. Sheffield: Sheffield Academic Press; 2001. p. 59–66.

43. Brook AH. Multilevel complex interactions between genetic, epigenetic and environmental factors in the aetiology of anomalies of dental development. Archives of Oral Biology. 2009;54(1):S3–S17.

44. Brook AH, Brook O’ Donnell M, Hone A, Hart E, Hughes TE, Smith RN, et al. General and craniofacial development are complex adaptive processes influenced by diversity. Australian Dental Journal. 2014;59(1):13–22.

45. Brook AH, Koh KSB, Toh VKL. Influences in a biologically complex adaptive system: Environmental stress affects dental development in a group of Romano-Britons. International Journal of Design and Nature and Ecodynamics. 2016;11(1):33–40.

46. Nanci A. Ten Cate’s Oral Histology : Development, Structure, and Function. Saint Louis: Saint Louis: Elsevier; 2012.

47. Brickley M. Cribra orbitalia and porotic hyperostosis: A biological approach to diagnosis. American journal of physical anthropology. 2018;167(4):896.

48. Godde K, Hens SM. An epidemiological approach to the analysis of cribra orbitalia as an indicator of health status and mortality in medieval and post-medieval London under a model of parasitic infection. American Journal of Physical Anthropology. 2021;174(4):631–45.

49. Walker PL, Bathurst RR, Richman R, Gjerdrum T, Andrushko VA. The causes of porotic hyperostosis and cribra orbitalia: A reappraisal of the iron-deficiency-anemia hypothesis. American Journal of Physical Anthropology. 2009;139(2):109–25.

50. Baker BJ, Crane-Kramer G, Dee MW, Gregoricka LA, Henneberg M, Lee C, et al. Advancing the understanding of treponemal disease in the past and present. American Journal of Physical Anthropology. 2020;171(S70):5–41.

51. Armelagos GJ, Goodman AH, Harper KN, Blakey ML. Enamel hypoplasia and early mortality: Bioarcheological support for the Barker hypothesis. Evolutionary Anthropology: Issues, News, and Reviews. 2009;18(6):261–71.

52. Brook AH, Smith JM. The Aetiology of Developmental Defects of Enamel: A Prevalence and Family Study in East London, U.K. Connective Tissue Research. 1998;39(1-3):151–6.

53. Brook AH, Smith JM. Hypoplastic enamel defects and environmental stress in a homogeneous Romano-British population. Eur J Oral Sci. 2006;114:370–4.

54. Halcrow SE, Tayles N. Stress near the start of life? Localised enamel hypoplasia of the primary canine in late prehistoric mainland Southeast Asia. Journal of archaeological science. 2008;35(8):2215–22.

55. Hillson S, Antoine D. The mechanisms that produce the defects of enamel hypoplasia. Am J Phys Anthropol. 2011;144(52):163–.

56. Beaumont J, Atkins EC, Buckberry J, Haydock H, Horne P, Howcroft R, et al. Comparing apples and oranges: Why infant bone collagen may not reflect dietary intake in the same way as dentine collagen. American Journal of Physical Anthropology. 2018;167(3):524–40.

57. Brook A, Fearne J, Smith J. Environmental causes of enamel defects. Dental Enamel. 1997;205:212–25.

58. Brickley MB, Kahlon B, D’Ortenzio L. Using teeth as tools: Investigating the mother-infant dyad and developmental origins of health and disease hypothesis using vitamin D deficiency. American journal of physical anthropology. 2019.

59. D’Ortenzio L, Kahlon B, Peacock T, Salahuddin H, Brickley M. The rachitic tooth: Refining the use of interglobular dentine in diagnosing vitamin D deficiency. International Journal of Paleopathology. 2018;22:101–8.

60. Mellanby M. The influence of diet on the structure of teeth: May Mellanby, Physiol. Rev., October, 1928, viii, 4. International Journal of Orthodontia, Oral Surgery and Radiography. 1929;15(4):388–90.

61. Mellanby M. Diet and the Teeth: An Experimental Study. Part III. The Effect of Diet on Dental Structure and Disease in Man. Journal of the American Medical Association. 1934;102(18):1523–.

62. Nikiforuk G, Fraser D. Etiology of Enamel Hypoplasia and Interglobular Dentin: The Roles of Hypocalcemia and Hypophosphatemia. Metabolic Bone Disease and Related Research. 1979;2(1):17–23.

63. Beckett RG. Paleoimaging: a review of applications and challenges. Forensic science, medicine, and pathology. 2014;10(3):423–36.

64. Beckett RG, Conlogue GJ, Nelson A. Case Studies for Advances in Paleoimaging and Other Non-Clinical Applications. Milton: Taylor and Francis; 2020.

65. Bruker. Position, Scan, Reconstruct and Analyze: Bruker; 2022 [Available from: https://www.bruker.com/en/products-and-solutions/microscopes/3d-x-ray-microscopes/skyscan-1272.html.

